# SARS-CoV-2 containment was achievable during the early stage of the pandemic: a retrospective modelling study of the Xinfadi outbreak in Beijing

**DOI:** 10.1101/2022.09.12.22279850

**Authors:** Yan Wang, Kaiyuan Sun, Yang Pan, Lan Yi, Da Huo, Yanpeng Wu, Shuaibing Dong, Jinxin Guo, Xiangfeng Dou, Wei Wang, Shuangsheng Wu, Xufang Bai, Hongjie Yu, Quanyi Wang

## Abstract

Prior to the emergence of the Omicron variant, many cities in China had been able to maintain a “Zero-COVID” policy. They were able to achieve this without blanket city-wide lockdown and through widespread testing and an extensive set of nonpharmaceutical interventions (NPIs), such as mask wearing, contact tracing, and social distancing. We wanted to examine the effectiveness of such a policy in containing SARS-CoV-2 in the early stage of the pandemic. Therefore, we developed a fully stochastic, spatially structured, agent-based model of SARS-CoV-2 ancestral strain and reconstructed the Beijing Xinfadi outbreak through computational simulations. We found that screening for symptoms and among high-risk populations served as methods to discover cryptic community transmission in the early stage of the outbreak. Effective contact tracing could greatly reduce transmission. Targeted community lockdown and temporal mobility restriction could slow down the spatial spread of the virus, with much less of the population being affected. Population-wide mass testing could further improve the speed at which the outbreak is contained. Our analysis suggests that the containment of SARS-CoV-2 ancestral strains was certainly possible. Outbreak suppression and containment at the beginning of the pandemic, before the virus had the opportunity to undergo extensive adaptive evolution with increasing fitness in the human population, could be much more cost-effective in averting the overall pandemic disease burden and socioeconomic cost.

## Introduction

Three years into the coronavirus disease 2019 (COVID-19) pandemic, we have witnessed the remarkable adaptive evolution of SARS-CoV-2 as it has spread and infected vast numbers of people across the globe. Numerous variants of concern, including Alpha, Beta, Gamma, Delta, and Omicron, have emerged to achieve global/regional dominance with increasing transmissibility and/or immune escape properties[1], rapidly replacing previously circulating variants and causing multiple epidemic waves. Phylogenetic analysis of the SARS-CoV-2 genome suggests that spike S1 has been the focus of adaptive evolution, with a high ratio of nonsynonymous to synonymous divergences 2.5 times greater than that of the influenza haemagglutinin HA1, than at the beginning of the 2009 H1N1 pandemic[2]. In addition, there is no evidence suggesting that SARS-CoV-2 is evolving in the direction of attenuated severity, with the Alpha and Delta variants acquiring increased severity[3] compared to the ancestral strain, while the Omicron variants may have lower severity as compared to Delta[4, 5]. Despite the development of highly effective SARS-CoV-2 vaccines and therapeutics at record speeds, the COVID-19 pandemic has caused more than 577 million infections and 6 million deaths worldwide as of Aug 4, 2022[6], while some countries including China have yet to experience large-scale epidemics that affect the majority of the population. Currently, the adaptive evolution of SARS-CoV-2 continues with no sign of slowing down, with multiple sublineages of Omicron emerging continuously (Figure 1A) and causing recurring Omicron waves despite high levels of population immunity achieved by vaccination and natural infection[7]. In contrast, at the beginning of the pandemic, even with neglectable population immunity (i.e., no effective vaccine available), due to the limited transmissibility of the ancestral strain (*R*_*0*_=2.5[8]), multiple regions across all socioeconomic status successfully achieved temporal suppression of SARS-CoV-2 in the early stage of the pandemic through the implementation of nonpharmaceutical interventions (NPIs) (Figure 1B). In retrospect, had the virus been successfully contained in the early stage of the pandemic, a great deal of the global morbidity, mortality and tremendous socioeconomic costs could have been avoided.

**Figure. 1.**
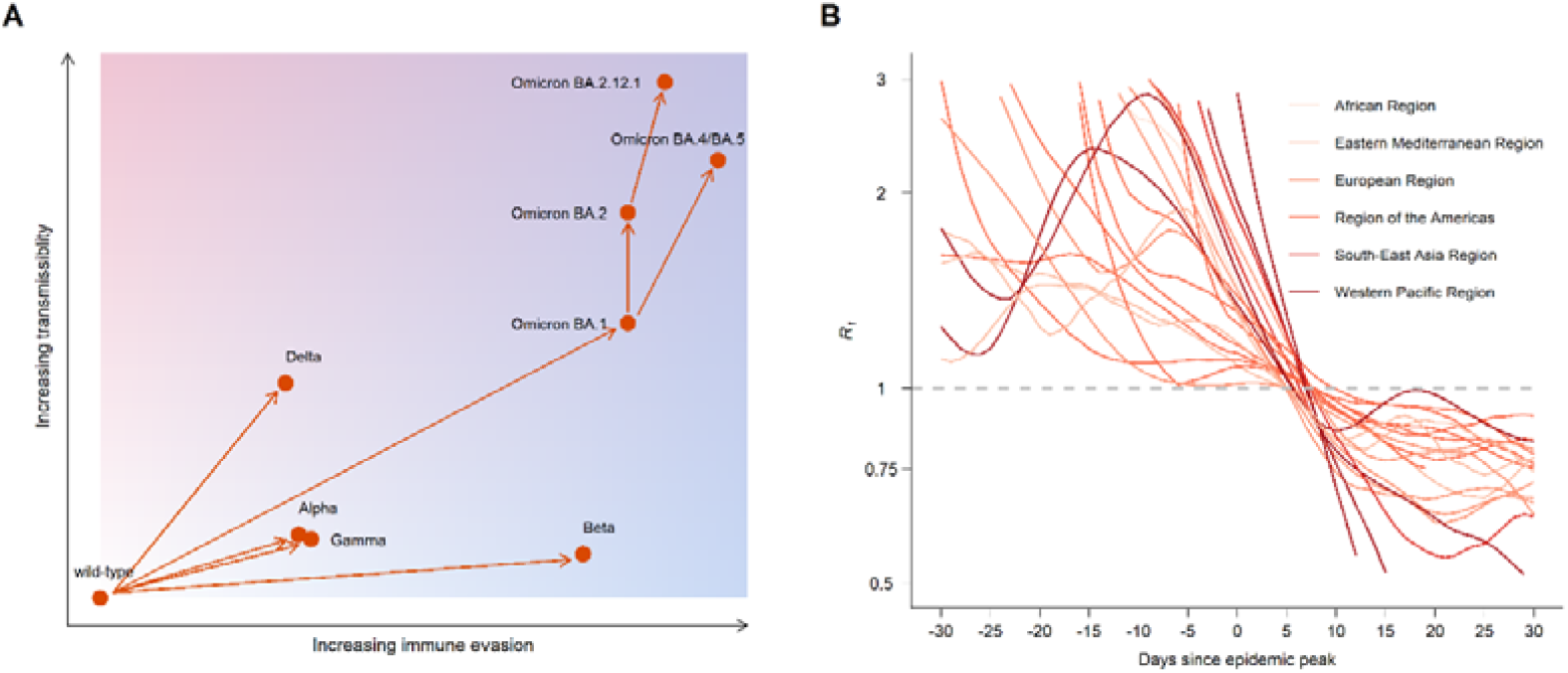
Increasing transmissibility and immune evasion properties of SARS-CoV-2 variants and the controllability of the SARS-CoV-2 ancestral strain. **A**. Schematic plot demonstrating the evolutionary trajectory of SARS-CoV-2 variants along the dimensions of transmissibility and immune evasion properties (detailed in Supplementary Materials, p24). The solid lines and the arrows indicate the mutation direction of the virus. **B**. Time-varying effective reproduction numbers (*R*_*t*_) during the first epidemic wave caused by the ancestral strain in different countries/states (detailed in Supplementary Materials, p24-26). All *R*_*t*_ time series were aligned by the epidemic peak. The horizontal dashed line indicates the epidemic threshold. The data sources and countries/states of each region are listed in Supplementary Table S7.

In this study, we conducted a retrospective analysis of a local SARS-CoV-2 containment against the ancestral strain to quantitatively assess the feasibility and conditions of containing the virus before it had the opportunity to accumulate its highly transmissive and immune-evasive properties. Specifically, we analysed the highly detailed epidemiological and contact tracing data of the Beijing Xinfadi outbreak in China that lasted from June 11 to July 10, 2020. The outbreak was caused by the ancestral strain of SARS-CoV-2, with the D614G mutation reintroduced from outside China[9] shortly after the successful suppression of the initial wave in Wuhan that ended in March 2020[10]. The Xinfadi outbreak is ideal to mimic a generic scenario of initial containment for countries outside China following the emergence and exportation of SARS-CoV-2 ancestral strain, as few adaptive mutations had accumulated at that time and most epidemiological features (i.e., the transmissibility, infectivity and pathogenicity) remaining intact. Population immunity was also negligible with no available vaccine and few prior SARS-CoV-2 infections in the population of Beijing during the period of the Xinfadi outbreak. The timing gap between the initial and the Xinfadi outbreaks also allowed Beijing to expand its SARS-CoV-2 molecular testing capacity, permitting mass testing of the population at risk of infection. As a result, the Xinfadi outbreak did not cause a city-wide lockdown by adopting more targeted interventions supported by a population-wide mass testing programme. We developed a spatially structured, individual-based model of SARS-CoV-2 transmission and nonpharmaceutical interventions to reconstruct the spread and containment of the Xinfadi outbreak in silico. The model was informed by high-resolution population mobility data and allowed us to explicitly model the targeted testing and intervention programme at high spatial resolution. Using the model, we demonstrated that a multilayer deployment of contact tracing, case isolation/contact quarantine, targeted lockdown, and mobility restriction could easily contain outbreaks caused by the SARS-CoV-2 ancestral strain, even without the requirement of blanket mass testing or city-wide lockdowns. More broadly, our study suggests that the critical opportunity window for containing a newly emerged pathogen is in the very early stage of the pandemic, when the pathogen’s adaptation in the human host may be far from optimal. Once a pathogen has spread across a population in large numbers, adaptive evolution could lead to the emergence of variants with enhanced transmissibility and immune evasion, increasing the difficulty of achieving containment. In addition, unmitigated spreading could also lead to spillover into other animal reservoirs[11-14], leaving pathogen eradication impossible to achieve.

## Methods

### Overview

Here, we conducted a modelling study of the Beijing Xinfadi outbreak to evaluate the feasibility and conditions of achieving SARS-CoV-2 containment through a comprehensive set of targeted interventions instead of city-wide lockdowns. In particular, we first analysed the detailed line list of SARS-CoV-2 case data to provide a quantitative description of the outbreak. Next, we developed a fully stochastic, spatially structured agent-based model to reconstruct the containment effort and recover the epidemiologic patterns observed in the epidemiological data. Last, we created 8 different hypothetical scenarios at 8 different levels of intervention intensities to dissect the relative contribution of each intervention individually to the overall containment of the Xinfadi outbreak.

### Data

Individual-level line list data recording the demographics, locations, exposures, symptoms and epidemiological links of the 368 SARS-CoV-2-infected individuals, as well as their 7,443 close contacts, were compiled by the Beijing Municipal Center for Disease Control and Prevention. Infections were identified between June 11 and July 10, 2020, during the Xinfadi outbreak. High-resolution mobile phone data, provided by the major mobile phone carrier in Beijing, were aggregated to describe the population mobility patterns in Beijing at the street/town level (Supplementary Materials, p3).

### Statistical analysis

We provide descriptive statistics of the outbreak, including spatiotemporal distributions and age profiles of the infected individuals. The distribution of the serial interval (time interval between onset of symptoms of the primary case and of their secondary cases) and the infectiousness profile (transmission probability from the primary case to the secondary cases over time) were estimated through probabilistically reconstructing infector-infectee transmission chains (Supplementary Materials, p3-5). The population flows (daily number of observed trips between streets/towns in Beijing) before and during the outbreak were also described based on mobility data, assuming that the mobility patterns of mobile phone users were representative of the general population.

## Model structure

### Initial infections

We initialized the model through the seeding of 169 infected workers at the Xinfadi Market, assuming that all subsequent infections outside the market were later generations of them. For each initial infection *i*, based on the recorded timing of symptom onset/diagnosis, the time of infection 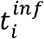 was stochastically sampled by randomly drawing either from the incubation period distribution *P*_*incu*_(*τ*) (for symptomatic infection) or the distribution of the time delay from infection to laboratory diagnosis *P*_*diag*_(*τ*) (for asymptomatic infection), conditional on their exposure window (Supplementary Materials, p5-6).

### Epidemiological features

For unmitigated transmission, we assumed that the basic reproduction number (*R*_0_) of the SARS-CoV-2 ancestral strain was 2.5[8]. We assumed that the generation interval followed a gamma distribution with a mean of 6.7 days and a standard deviation of 1.8 days[15]. The incubation period, derived from the epidemiological data, was assumed to follow a gamma distribution with a mean of 5.8 days and a standard deviation of 3.8 days. We hypothesized that the proportion of symptomatic infections *Φ*_*symp*_ increased with age, with 53.3%, 67.9% and 80.3% of infected individuals (without vaccination) who developed symptoms belonging to the 0-18 years, 19-59 years and 60 or older age groups, respectively[16].

### SARS-CoV-2 transmission as branching processes

To simulate the transmission of SARS-CoV-2 at the individual level, in the absence of NPIs, we assigned initial infection *i*’s reproduction number *R*_*i*_ (number of secondary infections caused by *i*) by drawing from a negative binomial distribution NB(*R*_0_, k), where the mean of the negative binomial distribution *R*_0_ is the basic reproduction number (population average of *R*_*i*_), and *k* is the dispersion parameter capturing the heterogeneity of SARS-CoV-2 transmission. Therefore, individual *i* would cause a total of *R*_i_ secondary infections. We assumed that the shape of each infected individual’s infectiousness profile followed the distribution of the generation interval. Thus, the timing of transmission *τ*_*ij*_ from individual i to individual *j* ∈ {*R*_*i*_} is given by 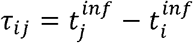, where *τ*_*ij*_ is drawn from the generation interval distribution *P*_*GI*_ (*τ*). For *j* (and all subsequent infections), the infection episode could be assigned as either symptomatic or asymptomatic based on the age-specific asymptomatic rate of SARS-CoV-2 *Φ*_*asymp*_. For symptomatic infection episodes, we assigned his or her time delay from infection to symptom onset τ_incu_ by drawing from the incubation period distribution *P*_*incu*_(*τ*) (Supplementary Materials, p6-8).

### Population structure

We divided the population into general workers, service workers and nonworkers. Service workers were further divided into workers at the Xinfadi Market and other service workers. When transmission between primary infection i and secondary infection *j* occurred, we first generated the setting of the transmission event (i.e., at home, in the workplace, or in the community) permitted by the occupation of the primary infection i. School transmission was not considered in this study because most schools and colleges did not reopen at the time of the outbreak. For any transmission event that occurs in the community, the activity of primary infection i (Act_i_) and secondary infection *j* (Act_j_) at the time of transmission was assigned based on their occupation. The age of secondary infection individual *j* was then assigned based on transmission setting, age-stratified contact matrices and age-specific susceptibility to SARS-CoV-2 infection. We further probabilistically generated J’s sex and determined J’s occupation in accordance with his or her age, conditional on the transmission setting, activity and i’s occupation (Supplementary Materials, p8-12).

### Population interaction network based on spatially resolved mobility patterns

Locations of secondary infections were stochastically allocated using a spatially structured network interaction model. We started with the initial infected individuals, whose home addresses and workplaces (i.e., the Xinfadi Market in Huaxiang Street, Fengtai District, Beijing) were collected through epidemiological investigations. We layered their home addresses into 3 levels with incremental spatial accuracy: i) the street/town (the basic-level administrative divisions in China); ii) the block; and iii) the household. We created a unique block identification number *blk_id* for each residential block and a unique household identification number *hh_id* for each family, where residents of a block shared the same *blk_id* and members of a household shared the same *hh_id*. Then, by developing a network interaction model, we probabilistically generated the residential locations (i.e., the street/town, *blk_id* and *hh_id*), the workplace (if he or she worked), and the locations of community activities (if the transmission events occurred through social activities) for each secondary infection based on the transmission setting and the spatially resolved mobility patterns in Beijing (Supplementary Materials, p12-19).

### Nonpharmaceutical interventions (NPIs)

The details of the intervention measures in response to the Xinfadi outbreak have been previously described[17, 18]. We categorized these measures into eight types: (i) symptom-based surveillance in health care facilities and communities; (ii) mask-wearing order in public places; (iii) closure of the Xinfadi Market; (iv) quarantine and testing of key populations associated with the Xinfadi Market; (v) systematic tracing, quarantine and testing of close contacts; (vi) confinement of residential communities with detected infections; (vii) population mobility restrictions based on the regional risk levels and (viii) mass testing at streets/towns with detected infections. A brief description of each NPI is given below. Details of the implementations of the simulation can be found in Supplementary Materials (p19-24).

- **Symptom surveillance:** Individuals who presented symptoms consistent with the clinical presentation of SARS-CoV-2 during health care consultations at hospitals and local clinics were considered SARS-CoV-2 suspected cases and were provisionally isolated in designated facilities. At least 3 subsequent RT□PCR tests for SARS-CoV-2 diagnosis were conducted on the 1^st^, 3^rd^ and 7^th^ days of isolation. If the suspected case was diagnosed by molecular tests, they remained in the hospital and were treated until they were fully recovered and no longer infectious. After the Xinfadi Market outbreak, to heighten case detection through symptom surveillance, routine temperature checking was implemented in public spaces, such as workplaces, markets, shopping malls, subway stations, railway stations and airports. Any individual with suspected symptoms of SARS-CoV-2 was asked to seek medical attention.
- **Mask wearing:** Mask wearing was required in public spaces, including hospitals, public transportation, markets, shopping malls, and entertainment venues.
- **Closure of the Xinfadi Market:** The Xinfadi Market was immediately closed on June 13, 2020, after the first positive sample was detected in the market.
- **Quarantine and testing of key populations:** During the Xinfadi outbreak, individuals associated with the Xinfadi Market were defined as the key population, including workers at the Xinfadi Market, visitors to the Xinfadi Market and residents living around the Xinfadi Market. Workers at the Xinfadi
- Market were assessed to be at the highest risk. They were immediately quarantined in centralized facilities for medical observation for at least 14 days. Visitors who had been to the Xinfadi Market between May 30 and June 12, 2020, were assessed as the subhigh-risk group, and a 14-day home quarantine was needed. Residents living around the Xinfadi Market, with moderate exposure risk, were confined to their living communities for at least 14 days. Enhanced RT___PCR testing was conducted on key populations during quarantine or confinement.
- **Contact tracing:** Close contact was defined as a person who interacted with a person with a confirmed or suspected COVID-19 case from 4 days before to 14 days after illness onset or with an asymptomatic carrier from 4 days before to 14 days after collection of the first positive sample. Close contacts were further grouped into household contacts, work contacts and community contacts based on their transmission setting. Epidemiological investigations to identify close contacts, including manual investigations (e.g., phone calls, interviews) and electronic tracing (e.g., mobile apps, online databases), were completed within 24 hours. Centralized quarantine for at least 14 days was required for all close contacts, with periodic RT□PCR testing at the 1^st^, 4^th^, 7^th^ and 14^th^ days of quarantine and the 2^nd^ and 7^th^ days after discharge.
- **Residential community confinement:** The residential communities with detected infections were on lockdown at the block level until 14 days after the identification of the last case, with stay-at-home orders for all residents other than essential workers. Supplies of living necessities were provided by community workers.
- **Mobility restrictions:** Unrestricted movement was allowed in low-risk areas, i.e., streets/towns without or with one detected infection. The street/town was upgraded to moderate risk once it had reported more than one infection, with entertainment venues being closed and mass gatherings being prohibited. Residents in moderate-risk areas were required to avoid unnecessary travel. Streets/towns with more than 5 infections were upgraded to high-risk status with more stringent population mobility restrictions being implemented. The street/town was downgraded to low risk if no new infections were reported for 14 consecutive days, with mobility restrictions being gradually lifted.
- **Mass testing:** Multiround mass testing at the street/town level was immediately activated and usually completed within 3-4 days when a new infection was reported. A 3:1 or 5:1 pooled sample approach was used to expand the RT□PCR capacity and increase cost-effectiveness.

### Simulation

Starting with the 169 infected workers at the Xinfadi Market (i.e., initial infections), we first simulated the transmission chain in the absence of NPIs based on the branching process model. We simulated the transmission chain until reaching July 10, 2020, i.e., the end date of the Xinfadi outbreak. We then created eight intervention scenarios, Level 1 to Level 8, by progressively layering additional NPIs on top of the prior scenario:

- Level 1: only symptom surveillance;
- Level 2: Level 1 + mask wearing;
- Level 3: Level 2 + closure of the Xinfadi Market;
- Level 4: Level 3 + quarantine and testing of key populations;
- Level 5: Level 4 + contact tracing;
- Level 6: Level 5 + community lockdowns;
- Level 7: Level 6 + mobility restrictions; and
- Level 8: Level 7 + mass testing.

We simulated the effect of NPIs by pruning the unmitigated chains of transmission by removing branches that would otherwise be interrupted by the corresponding NPIs of the scenario of interest. We ran 500 simulations for each scenario to capture the stochasticity of the transmission process. For the Level 8 intervention scheme (corresponding to the NPIs adopted to contain the Xinfadi outbreak), we summarized the following characteristics of each simulation compared with the observed epidemiological patterns to test the model performance: i) daily number of detected infections by clinical severity; ii) number of detected infections of each street/town; iii) number of detected infections of each age group; iv) serial interval; and v) infectiousness profile since onset of symptoms. Finally, for each simulation, the following summary statistics were calculated to quantify the impact of each intervention individually: i) the overall effective reproduction number (*R*_*eff*_), defined as the average of the individual reproduction number of each individual infected after the implementation of NPIs; ii) the total number of infections; and iii) the proportion of undetected infections. The branching process model was coded in Python 3.10. The statistical analyses and visualization were performed using R software, version 4.0.2.

## Results

The Beijing Xinfadi outbreak was initially identified on June 11, 2020, through a suspected case with SARS-CoV-2 symptoms at a local fever clinic. After initial laboratory confirmation and contact tracing investigation, the Xinfadi wholesale market located in a suburb of Beijing was identified as the most likely source of exposure, triggering swift market closure and isolation followed by complete screening of market workers using reverse transcription polymerase chain reaction (RT-PCR) testing. A total of 169 market workers tested positive with a clear clustering pattern, triggering an emergency outbreak investigation and expansive contact tracing across the city among people who had a direct epidemiologic link with the identified infected individuals and/or had visited the Xinfadi Market up to two weeks prior to the initial identification of the outbreak. On average, 20 contacts per infection were traced throughout the Xinfadi outbreak. Supplementing the contact tracing effort, starting from June 13, 2020, city-wide mass testing was implemented to uncover cryptic transmission in the community and bridge unconnected transmission chains with an initial testing capacity of 100,000 tests per day ramping up to 500,000 tests per day by July 7, 2020. In total, 11 million people were screened, representing 50% of the total population in Beijing, of which more than 550,000 individuals with high exposure risk, including close contacts and key populations associated with the Xinfadi Market, were screened more than once. Targeted lockdowns at the blocks with positive cases (with 500-230,000 residents per block compared to 21.9 million people for the city of Beijing) were implemented to stop onward community transmission. Additional mobility reductions (temporarily reducing the mobility of individuals based on the risk level of the corresponding street/town, rather than city-wide lockdowns) were put in place to slow down the spatial dissemination of the outbreak.

A total of 368 SARS-CoV-2 infections were reported during the Xinfadi outbreak in Beijing, China, of which 335 (91.03%) were identified as confirmed cases and the other 33 (8.97%) were identified as asymptomatic infections (Figure 2A). The spatial distribution of the infections varied substantially by administrative divisions, with most of the infections clustered in or around Huaxiang Street, where the Xinfadi Market is located (Figure 2B). Overall, most infections were in the working age population, aged between 20 and 59 years, with only 3.3% of the infections occurring among people aged under 18 years and 9% occurring among people aged 60 years or older (Figure 2C). To estimate the duration of the serial interval, we analysed 34 transmission pairs with available symptom onset dates. We found that the best fitting distribution of the serial interval was a Weibull distribution with a mean of 3.3 (median: 3.2, interquartile range (IQR): 0.7-5.8) days (Figure 2D). The estimated serial interval was shorter than the parameterized intrinsic generation interval of 6.7 days[15], suggesting that nonpharmaceutical interventions (NPIs) successfully blocked further transmission through case isolation and contact quarantine[19]. We estimated the proportion of presymptomatic transmission (area under the curve, Figure 2E) at 68.6%, with 95% of transmission events occurring before 3.9 days after symptom onset. As interventions, such as isolation of infected individuals and quarantine of contacts, can prevent potential infectors from contacting susceptible individuals and thus effectively shorten the infectious period, it is important to stress that our estimates reflect the impact of NPIs.

**Figure. 2.**
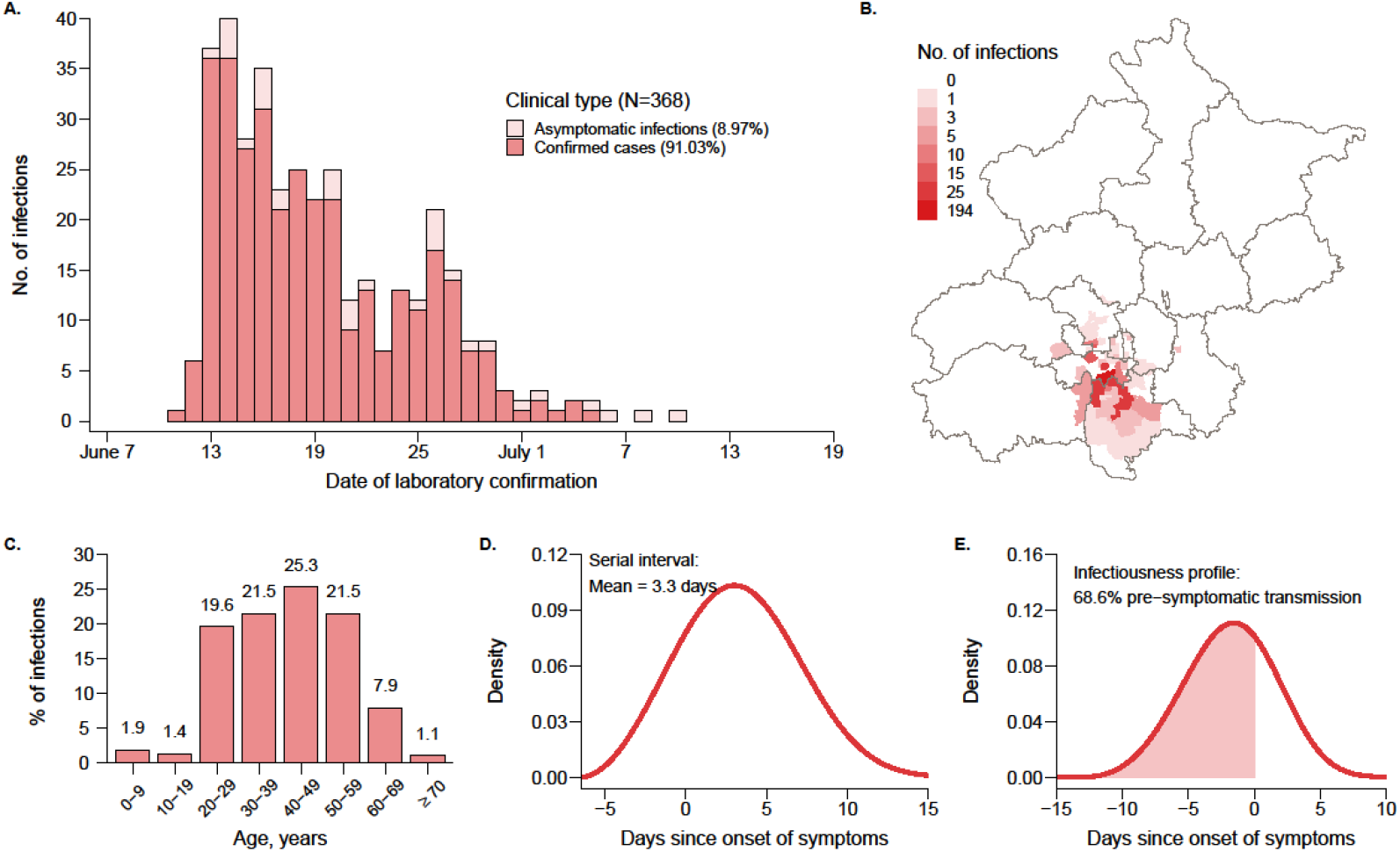
The epidemiologic patterns observed during the Xinfadi outbreak. **A**. Temporal distribution of SARS-CoV-2 infections stratified by clinical type. **B**. Spatial distribution of SARS-CoV-2 infections. **C**. Age distribution of SARS-CoV-2 infections. **D**. Estimated distribution of the serial interval. **E**. Estimated distribution of the infectiousness profile.

Next, we built a stochastic, spatially structured, agent-based model of both SARS-CoV-2 infection and NPIs to reconstruct the Beijing Xinfadi outbreak in silico. We ran the model under the same initial condition with 500 repeat simulations to capture the stochasticity of the transmission process. Simulations that eventually lead to outbreaks closely resemble those observed in the real world. In Figure 3A, we present one realization of the temporal distributions of the reported infections. Similar to the observed distribution in Figure 2A, a total of 355 infections were detected in the simulated outbreak, of which 18.87% were asymptomatic infections and 81.13% were confirmed cases. Figure 3B shows the average number of reported infections of each street/town, aggregating from the results of 500 simulated outbreaks, with most of the infections detected around the Xinfadi Market. The aggregated age profile also follows the observed distributions in Figure 2C, where most of the infections occurred in the working age population (Figure 3C). Based on the simulated transmission pairs, the mean realized serial interval was estimated to be 3.7 (median: 4.2, IQR: -0.8, 8.7) days (Figure 3D). Our estimate using the observed epidemiological data (Figure 2D) falls within the interquartile range. Similar to the observed infectiousness profile in Figure 2E, we found that most simulated transmission events occurred during the presymptomatic phase (62.6%), with 95% of transmission events occurring before 5.3 days after symptom onset (Figure 3E).

**Figure. 3.**
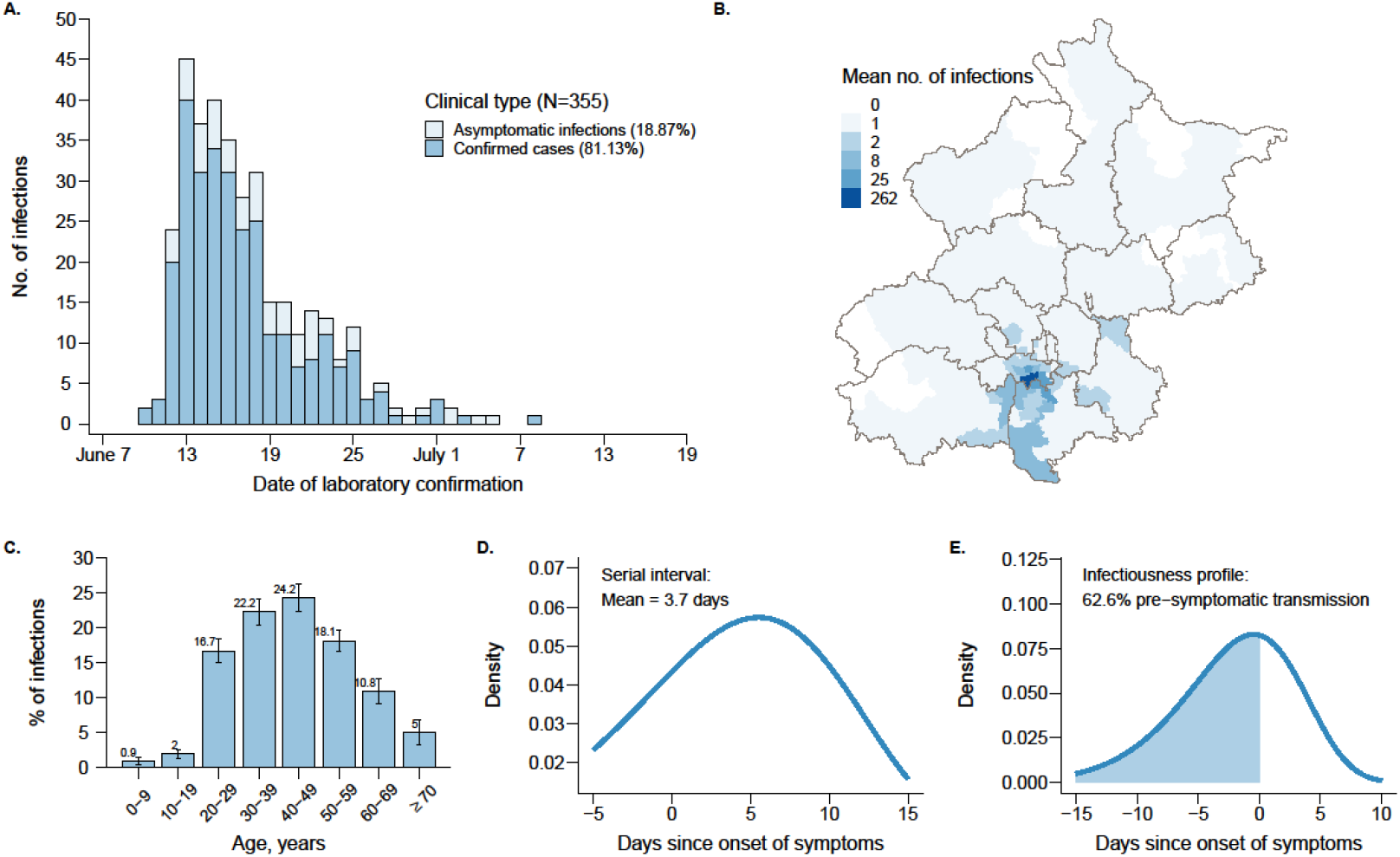
The epidemiologic patterns simulated by reconstructing the containment efforts during the Xinfadi outbreak. **A**. Temporal distribution of SARS-CoV-2 infections stratified by clinical type. **B**. Spatial distribution of SARS-CoV-2 infections. **C**. Age distribution of SARS-CoV-2 infections. **D**. Estimated distribution of the serial interval. **E**. Estimated distribution of the infectiousness profile. Panel A represents the result of one simulated outbreak, while the results in Panels B-E are estimated based on 500 simulations.

Finally, to quantify the relative contribution of each individual NPI to outbreak containment, we consecutively added each intervention to the unmitigated chains of transmission. The estimated effective reproduction numbers (*R*_*eff*_) and total number of infections (*N*) before July 10 are reported in Figure 4A and Figure 4B. We found heterogeneity across simulations even under the same intervention intensity, reflecting the intrinsic stochasticity of SARS-CoV-2 transmission. We found that the outbreak could not be contained with only symptom surveillance (Level 1) due to the presymptomatic and asymptomatic transmission of SARS-CoV-2, with a median *R*_*eff*_ = 2.05 and a median *N* = 13421, respectively. Layering mask wearing (Level 2) and closure of Xinfadi Market (Level 3) did not lead to significant improvement, with all simulations having effective reproduction numbers larger than 1.8, well above the epidemic threshold. Quarantine of the key populations (Level 4) could remove the potential infections from the susceptible population at the early phase of viral shedding, thus effectively reducing the number of infections before July 10 (median *N* = 2967), but the onward transmission could not be suppressed (median *R*_*eff*_ = 1.78). Contact tracing (Level 5), in contrast, could significantly reduce onward transmission, leading to fewer infections (median *N* = 1141) and lower effective reproduction numbers (median *R*_*eff*_ = 1.19), but for most simulations, the estimated *R*_*eff*_ was still above the epidemic threshold, indicating that containment could not be achieved through Level 5 intervention intensity. With confinement of infected individuals’ residential communities (Level 6), the median *R*_*eff*_ hovered around the epidemic threshold of 1, resulting in highly stochastic outcomes with the probability of achieving containment at 50%. Implementing targeted population mobility restrictions (Level 7) would achieve the goal of containment, with the estimated *R*_*eff*_ ranging from 0.39 to 0.89 and the number of infections less than 1,000 for all simulations. Although the outbreak could be suppressed with Level 7 intervention intensity, additional mass testing at the street/town level was adopted during the Xinfadi outbreak, which could further reduce the effective reproduction number and the number of infections, with a median *R*_*eff*_ = 0.64 and a median *N* = 447, leading to faster clearance and fewer infections.

**Figure. 4.**
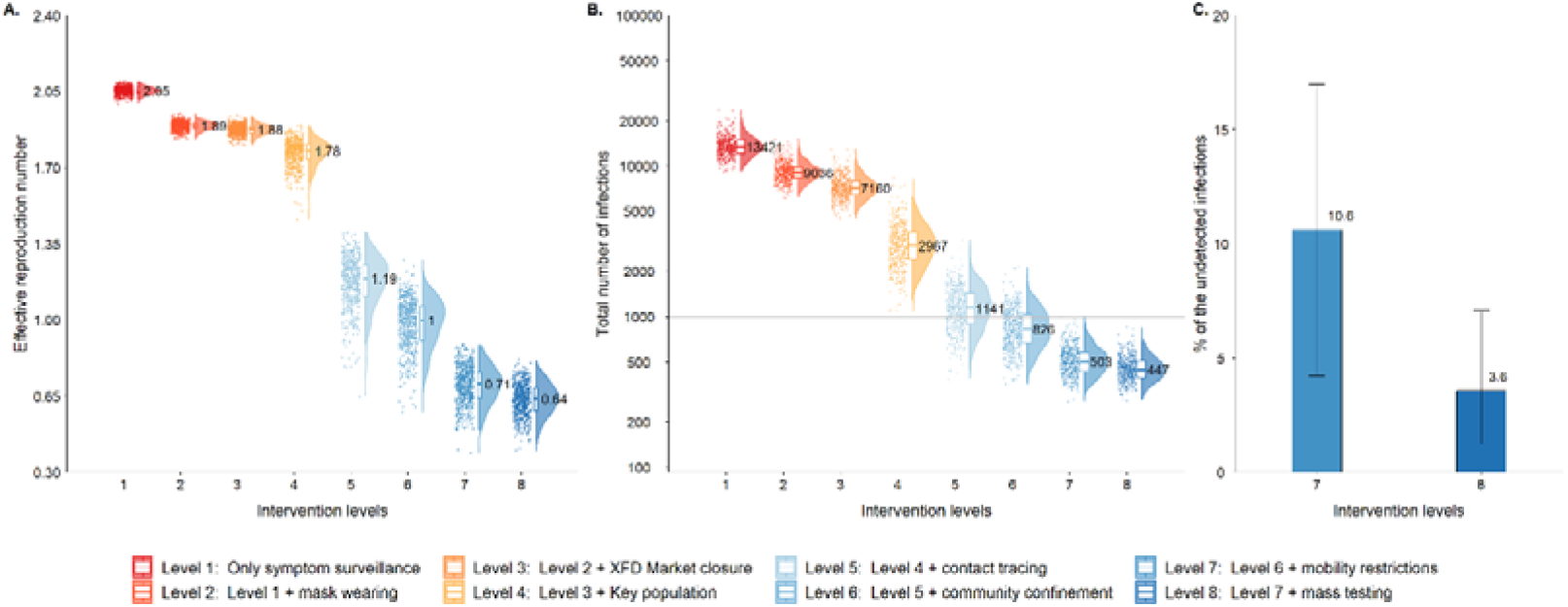
The impact of each NPI. **A**. Effective reproduction numbers under different intervention scenarios. **B**. Number of infections under different intervention scenarios. **C**. Proportion of undetected infections. The bars represent the median of 500 simulations, and the lines give the range of the 2.5 and 97.5 quantiles.

Further, we found that approximately 10.6% of the infections were undetected in the absence of mass testing (Level 7), while the number of undetected infections fell to 3.6% after the implementation of mass testing (Level 8), indicating that the outbreak could be contained earlier through mass testing, with more infections being detected and isolated, and thus the onward transmission could be truncated (Figure 4C).

## Discussion

In this study, we demonstrated through both empirical data and modelling analysis that the SARS-CoV-2 ancestral strain could have been contained through NPIs. We found that robust implementation of symptom surveillance and high-risk population screening served as sentinels to discover cryptic community transmission in the early stage of the outbreak. Early detection could help in reducing the overall demand of individual-based interventions, such as contact tracing and targeted community confinement, as the cost of such interventions increases proportionally to the number of active infections. Effective contact tracing combined with case isolation and close contact quarantine have been shown to substantially reduce transmission, highlighting the importance of training and maintaining epidemiologic teams with field experience. Targeted community lockdown at the block level, where new cases were discovered, could further limit undiscovered infections missed by contact tracing. This could prevent further spread as the transmission of SARS-CoV-2 demonstrates clear spatial clustering. In addition, targeted community confinement is more cost-effective than population-wide lockdowns, with fewer people being affected. Logistic support for maintaining the supplies of necessities is crucial to minimize the impact of societal disruption caused by confinement. Rapid turnaround of molecular testing for the confined population could help confirm and release infection-free individuals back to their normal lives as soon as possible. At the city level, temporal reductions in mobility (rather than blanket lockdowns) in and out of regions with high infection risk could slow down the spatial dissemination of the outbreak and speed up the viral eradication process. If conditions permit, population-wide mass testing could further improve the viral clearance speed. However, mass testing programmes are generally very costly, and the cost-effectiveness of such programmes needs to be taken into consideration. Our study clearly demonstrated that the containment of the SARS-CoV-2 ancestral strain would have been achievable through NPIs once a reliable and scalable diagnostic test became available. Real-world experience also suggests that multiple countries with different level of socioeconomic statuses have successfully achieved temporal eradication of SARS-CoV-2 in the early stages of the pandemic. For countries who did not choose containment, many were forced into blanket population lockdowns due to the high disease burden of mortality and morbidity as well as the risk of overwhelming the health care system but were still successful at suppressing the outbreak once strict measures were implemented (Figure 1B). This evidence suggests that a global containment of SARS-CoV-2 was certainly feasible at the beginning of the pandemic before the virus had the opportunity to evolve and adapt in the human population with greatly enhanced transmissibility and immune evasion properties, if all countries had decided to pursue such strategies collectively.

## Data Availability

All data produced in the present study are available upon reasonable request to the authors

## Ethical approval statement

Ethical approval and informed consent requirements were waived by the Institutional Review Board and Human Research Ethics Committee of the Beijing Center for Disease Prevention and Control (Beijing CDC) because this study was considered a continuation of the public health investigation associated with an emerging infectious disease.

## Acknowledgements

This study was supported by grants from the Key Program of the National Natural Science Foundation of China (82130093). The findings and conclusions in this study are those of the authors and do not necessarily represent the official position of the funding agencies, the National Institutes of Health, or the U.S. Department of Health ands Human Services.

## Author contributions

H.Y. and Q.W. designed and supervised the study. Y.P., D.H., S.D., X.D., and S.W. performed the field epidemiological investigations and collected the original data. Y. Wang, L.Y., Y. Wu, J.G., W.W., and X.B. helped with cleaning the original data. Y. Wang, K.S. and Y.P. carried out the development of the model, designed the simulations, performed statistical analysis and drafted the manuscript. L.Y. helped with constructing the figures. Q.W. and H.Y. performed a critical revision of the manuscript for important intellectual content. All authors were involved in the research and revision of the manuscript. All authors approved the final manuscript.

