## Supplementary materials for "SARS-CoV-2 containment was achievable during the early stage of the pandemic: a retrospective modelling study of the Xinfadi outbreak in Beijing"

### 1 Data sources

#### 1.1 Outbreak data

We collected individual records of 368 individuals with confirmed SARS-CoV-2 infections and their 7,443 close contacts in Beijing, China, from June 11 to July 10, 2020. The line list data, including individual-level demographics, geolocations, and epidemiological and clinical information, were extracted from the Notifiable Infectious Disease Reporting System and the Epidemiological Investigation Information System of the People’s Republic of China. Close contact data, including individual contact-level demographics, exposures and quarantine data, were collected from the Close Contacts Tracing and Management System.

#### 1.2 Mobility data

The aggregated mobile phone signalling data, describing the population mobility patterns in Beijing from June 1 to June 30, were provided by China Unicom, one of the leading mobile phone service providers in China, which holds the records of more than 12 million anonymous subscribers in Beijing. The data were aggregated as origin-destination matrices (O-D matrices) stratified by age group and hour, where the rows represent the origin streets/towns and the columns are the destination streets/towns. Each cell of the matrix represents the total number of trips of the subscribers from the origin to the destination in a given age group during a certain period of time. The O-D matrices were first constructed using the travel records of the subscribers of China Unicom and then rescaled to represent the mobility patterns of all mobile phone users based on the carrier penetration rate using the extrapolation algorithm provided by China Unicom.

### 2 Serial interval distribution and infectiousness profile

We estimated the distribution of the serial interval (time interval between onset of symptoms of the primary case and of their secondary cases) and the infectiousness profile (transmission probability from the primary case to the secondary cases over time) based on 1000 realizations of the reconstructed transmission chains, which were customized to the unique contact tracing data of the Xinfadi outbreak accounting for uncertainties in multiple transmission routes compatible with the observation.

Specifically, for each infection *i,* the infection time $t_{i}^{inf}$ was stochastically sampled by subtracting a randomly sampled incubation period $\tau_{i}^{incu}$(drawing from the incubation period distribution $P_{incu}\left( \tau\right)$) from the date of symptom onset $t_{i}^{sym}$. We assumed that the incubation period followed a gamma distribution with a mean of 5.8 days and a standard deviation of 3.9 days, which was estimated using the maximum likelihood method based on the individual records in the line list data.

The sampled time of infection $t_{i}^{inf}$ had to satisfy the following constraints:

- $t_{i}^{inf}$ had to fall within the start and end dates of the exposures identified by epidemiological investigation.
- For any infector-infectee pair, the infection time of infector $t_{infector}^{inf}$ had to be earlier than the infection time of infectee $t_{infectee}^{inf}$, i.e., $t_{infector}^{inf}< t_{infectee}^{inf}$.

For an infected individual *i* who had contact with multiple SARS-CoV-2-infected individuals, all other individuals in contact with *i* were potential infectors, except for those whom *i* infected. We first generated a series of possible infection dates for *i* conditional on his or her time of exposure to each infector, denoted as $t_{i}^{inf.1}$, $t_{i}^{inf.2}$, $t_{i}^{inf.3}\cdots$, and calculated the corresponding incubation period, denoted as $\tau_{i}^{incu.1}$, $\tau_{i}^{incu.2}$, $\tau_{i}^{incu.3}\cdots$. The probability that *k* was the source of infection of *i* was then calculated as $P\left( t_{i}^{inf}=t_{i}^{inf.k} \right)= P_{incu}\left( \tau=\tau_{i}^{incu.k} \right)$, where $k=1,2,3\cdots$. We chose the one with the highest probability as the source of infection of *i*.

The following individuals were excluded from the analysis:

- Individuals with sporadic infections that were not epidemiologically linked to other infections.
- Asymptomatic individuals.
- Individuals with symptomatic infections with missing onset dates.
- Individuals with symptomatic infections whose potential infectors had missing onset dates.

A total of 1000 realizations of transmission chains were stochastically reconstructed to account for uncertainties in both the timing and source of exposures. For all infector-infectee pairs in 1000 realizations of reconstructed transmission chains, we calculated the time interval between their dates of symptom onset (i.e., serial interval), as well as the time interval between symptom onset and transmission (i.e., for estimating the infectiousness profile over time). We fit three distributions (gamma, Weibull and lognormal) for both intervals using maximum likelihood, with shift parameters allowing negative values, and assessed the goodness of fit using the Akaike information criterion (AIC).

### 3 Model structure

#### 3.1 Initial infections

We defined the 169 infected workers at the Xinfadi Market as initial infections, assuming that all subsequent individuals outside the market were later infected by them. The demographics, locations, exposures, modes of detection, dates of symptom onset, and diagnostic information of initial infections were extracted from line list data. We created a unique household identification number ($hh\_id$) for each family, where members of a household shared the same $hh\_id$. For each household, the total number of family members ($hh\_size$) and number of infections included in the initial infections were further collected from close contact data. The time of infection $t_{i}^{inf}$ for each initial infection *i* was stochastically generated conditional on their exposure window. Specifically, among all 169 initial infections, 153 were symptomatic cases, and 16 were asymptomatic infections. Of the 153 symptomatic cases, 58 were diagnosed by computed tomography (CT) imaging followed by laboratory testing results, with clinical symptoms developing after hospitalization. For these cases, their symptom onset dates were not recorded in the line list data. We first imputed their symptom onset dates with their dates of initial CT scan (first time of imaging examination with CT infiltrates) plus a randomly selected time interval from the distribution of the time delay from initial CT scan to symptom onset (i.e., a normal distribution with a mean of 3.8 days and a standard deviation of 1.5 day)[1]. Then, for each symptomatic initial infection *i,* the time of infection $t_{i}^{inf}$ was sampled by randomly drawing from the incubation period distribution $P_{incu}\left( \tau\right)$ and subtracting this value from the reported time of symptom onset, $i.e., t_{i}^{inf}=t_{i}^{sym}-\tau_{i}^{incu}$, where $\tau_{i}^{incu}$ is the sampled incubation period and $t_{i}^{sym}$ is the date of symptom onset of *i*. Next, we fit the distribution of the time interval from infection to laboratory confirmation $P_{diag}\left( \tau\right)$ using the observed diagnosis dates and simulated infection time of all symptomatic cases and assumed that the same distribution was followed for asymptomatic infections. Therefore, we assigned the time of infection $t_{i}^{inf}$ for each asymptomatic infection by subtracting $\tau_{i}^{inf\_diag}$ from the time of laboratory confirmation $t_{i}^{diag}$, $i.e., t_{i}^{inf}=t_{i}^{diag}-\tau_{i}^{inf\_diag}$, where $\tau_{i}^{inf\_diag}$ is the sampled time delay from infection to diagnosis. The sampled time of infection $t_{i}^{inf}$ was constrained within the exposure window identified by epidemiological investigation.

#### 3.2 SARS-CoV-2 transmission as branching processes

- **Individual reproduction number:** We first generated each initial infection *i'*s reproduction number $R_{i}$ (number of secondary infections caused by $i$). For unmitigated transmission, we assumed that $R_{i}$ followed a negative binomial distribution $NB\left( R_{0}, k \right)$, where $R_{0}$ is the basic reproduction number representing the population average of $R_{i}$ and *k* is the dispersion parameter capturing the heterogeneity of SARS-CoV-2 transmission. For the Xinfadi outbreak (caused by the SARS-CoV-2 ancestral strain), we assumed the basic reproduction number *R_0_*=2.5[2] and dispersion parameter *k*=0.43[3].
- **Infection time:** For each secondary infection $j\in\left\{ R_{i} \right\}$, the infection time $t_{j}^{inf}$ is given by $t_{j}^{inf}=t_{i}^{inf}+\tau_{ij}$, where $\tau_{ij}$ denotes the generation interval of transmission from *i* to *j*, drawn from a gamma distribution, with a mean of 6.7 days and a standard deviation of 1.8 days[4].
- **Onset of symptoms:** Then, the infection episode of each individual $j$ could be assigned as either symptomatic or asymptomatic. We hypothesized that the proportion of individuals with symptomatic infection $\Phi_{symp}$increased with age, with 53.3%, 67.9% and 80.3% of infected individuals (without vaccination) developing symptoms belonging to the 0-18 years, 19-59 years and 60 or older age groups, respectively[5]. For a symptomatic infection episode, we assigned his or her symptom onset date $t_{j}^{sym}$ by drawing from the incubation period distribution $P_{incu}\left( \tau\right)$, $i.e., t_{j}^{sym}=t_{j}^{inf}+\tau_{j}^{incu}$, where $\tau_{j}^{incu}$ is the generated incubation period of individual $j$, following a gamma distribution with a mean of 5.8 days and standard deviation of 3.8 days, derived from the epidemiological data.

The parameters reflecting the transmission dynamics and the natural history of COVID-19 are summarized in Table S1.

**Table S1. Parameters reflecting the transmission dynamics and the natural history of COVID-19.**

| **Parameter** | **Description** | **Value** |
| --- | --- | --- |
| $R_{0}$ | Basic reproduction number | 2.5 |
| $k$ | Dispersion parameter capturing the heterogeneity of SARS-CoV-2 transmission | 0.43 |
| $P_{GI}(\tau)$ | Distribution of generation interval | Gamma distribution  (shape = 13.86, rate = 2.07) |
| $\Phi_{asymp}$ | Age-specific asymptomatic rate | 46.7% for 0-18 years  32.1% for 19-59 years  19.7% for 60+ years |
| $P_{incu}\left( \tau\right)$ | Distribution of the incubation period | Gamma distribution  (shape = 2.25, rate = 0.39) |

#### 3.3 Population structure reflecting demographics, transmission setting, and activity

- **Transmission setting:** When transmission between primary infection $i$ and secondary infection $j$ occurred, we first generated the transmission setting based on the predefined probability that the transmission event occurred at home ($\emptyset_{hh}$), in the workplace ($\emptyset_{wk}$) or in the community ($\emptyset_{cm}$). The hypothetical probabilities conditional on the occupation ($O_{i}$) and household size (${hh\_size}_{i}$) of primary infection $i$, as well as the timing of transmission from $i$ to $j$ ($t_{j}^{inf}$), are summarized in Table S2, where we assumed that i) all service workers worked in the community and ii) all general workers did not work on holidays or weekends.

**Table S2. Hypothetical probability that the transmission event occurred at home, in the workplace or in the community.**

| Condition | | Hypothetical probability | | |
| --- | --- | --- | --- | --- |
| $O_{i}$ | $t_{j}^{inf}$ | $\emptyset_{hh}$ | $\emptyset_{wk}$ | $\emptyset_{cm}$ |
| ${hh\_suscept}_{i}$ = 0^†^ |  |  |  |  |
| General worker | Workday | 0 | 0.6 | 0.4 |
|  | Holiday/weekend | 0 | 0 | 1 |
| Service worker | - | 0 | 0 | 1 |
| Nonworker | - | 0 | 0 | 1 |
| ${hh\_suscept}_{i}$ > 0 |  |  |  |  |
| General worker | Workday | 0.5 | 0.3 | 0.2 |
|  | Holiday/weekend | 0.7 | 0 | 0.3 |
| Service worker | - | 0.5 | 0 | 0.5 |
| Nonworker | - | 0.7 | 0 | 0.3 |

Note: ^†^ ${hh\_suscept}_{i}$ denotes the number of susceptible individuals in primary infection $i$’s home at the time of transmission. ${hh\_suscept}_{i}=0$ indicates that all the household members of primary infection $i$ have been infected before $t_{j}^{inf}$; thus, transmission from *i* to *j* could not occur at home, i.e., $\emptyset_{hh}$=0.

- **Activity in the community:** For any transmission event occurring in the community, the activity of primary infection $i$ ($\mathrm{Act}_{i}$) and secondary infection *j* ($\mathrm{Act}_{j}$) at the time of transmission was assigned based on their occupation. Service workers who worked in the community could be assigned either work or social activity, with probabilities of $\varphi_{cm}^{wk}$=0.6 and $\varphi_{cm}^{soc}$=0.4, respectively. For general workers and nonworkers, we assumed that only social activity was possible in the community (i.e., $\varphi_{cm}^{wk}=0$).
- **Age:** The age of secondary infection *j* was then assigned based on transmission setting, age-stratified contact matrices and age-specific susceptibility to SARS-CoV-2 infection. We defined the age-specific contact matrices as $C^{hh}$, $C^{wk}$ and $C^{cm}$ for household, workplace, and community contact, respectively. Each cell of the matrix ($c_{mk}^{T}$) represents the average number of contacts in age group *k* of an individual in age group *m* in a given setting *T*. For any contact in age group *k*, whether he or she became infected depended on the age-specific susceptibility to SARS-CoV-2 infection ${risk}_{k}^{inf}$. Therefore, the probability that *j* is in age group $a_{j}$ is given by

$$P_{a_{j}|a_{i}}^{T}=\frac{c_{a_{i}a_{j}}^{T} {risk}_{a_{j}}^{inf}}{\sum_{k} C_{a_{i}k}^{T} {risk}_{k}^{inf}}$$

where $a_{i}$ is the age group of primary infection *i.* The age-mixing contact matrices of the general population were derived from the contact survey data in China prior to the COVID-19 pandemic (Fig. S1)[6], while for workers at the Xinfadi Market (a special group of people, most of whom were working age living in the staff dormitories near the market), it was extracted from the close contact data of infected Xinfadi workers (Fig. S2). The age-specific susceptibility to SARS-CoV-2 infection, as presented in Table S3, was derived from a multivariate generalized linear mixed effect model (GLMM) based on 2,996 close contacts identified during the first epidemic wave (January-March 2020) in Beijing, China.

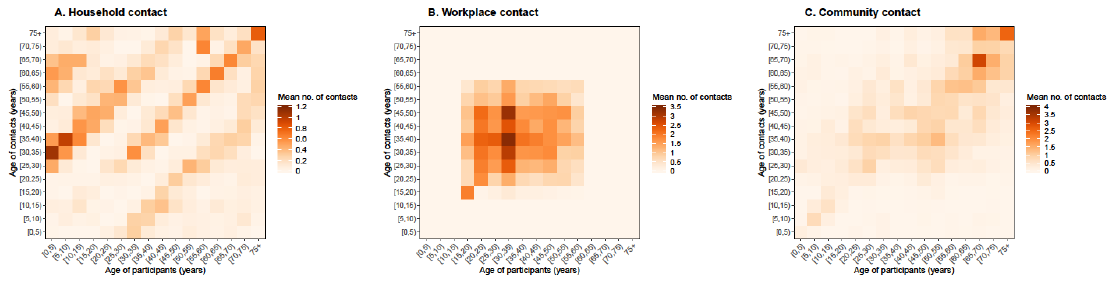

**Figure S1. Contact matrices of the general population by setting and age. A**. Contact matrix in households. **B**. Contact matrix in the workplace. **C**. Contact matrix in the community, where we assumed a 70% contact reduction among people aged 0-20 years due to the closure of off-campus training institutions in Beijing due to the COVID-19 pandemic. Each cell of the matrix represents the mean number of contacts that an individual in a given age group had, stratified by age groups. The colour intensity represents the number of contacts.

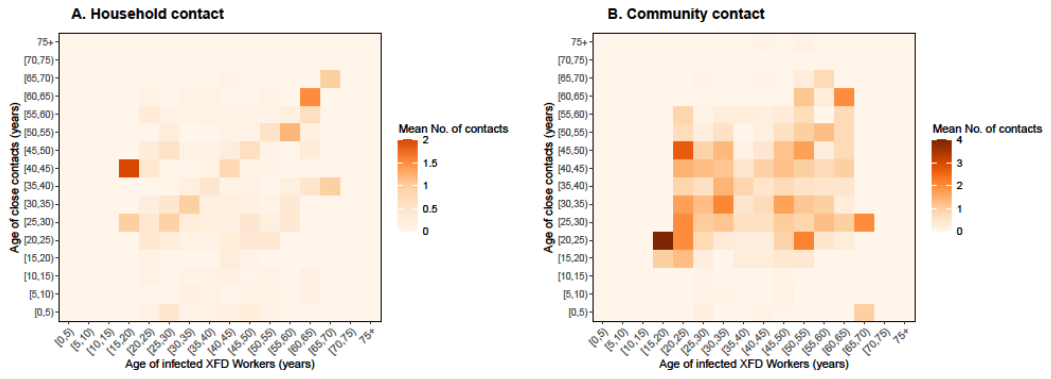

**Figure S2. Contact matrices of workers at the Xinfadi Market by setting and age. A**. Contact matrix in households. **B**. Contact matrix in the community.

**Table S3. Age-specific susceptibility to SARS-CoV-2 infection.**

| **Age group**  (*k*) | **Relative risk**  $({risk}_{k}^{inf})$ |
| --- | --- |
| 0-14 years | 0.25 |
| 15-29 years | 0.79 |
| 30-49 years | 1 |
| 50-64 years | 1.13 |
| 65+ years | 1.32 |

- **Sex:** The sex of secondary infections was generated completely at random, except for individuals infected in the workplace aged 55-59 years (who were males because females retire from work at 55 years of age).
- **Occupation:** To assign secondary infection $j$’s occupation, we first determined whether he or she was a worker (i.e., 18-59 years for a male; 18-54 years for a female) according to his or her age and sex. We then divided workers into general workers (GW) and service workers (SW). Service workers were further divided into workers at the Xinfadi Market (XFD-SW) and other service workers (Other-SW). For any infected worker $j$, we stochastically determined his or her specific occupation based on the predefined probability that the secondary infection $j$ worked as a general worker ($\psi_{j}^{GW}$), a service worker at the Xinfadi Market ($\psi_{j}^{XFD-SW}$) or a service worker in other places ($\psi_{j}^{Other-SW}$). The hypothetical probabilities conditional on the transmission setting ($T_{ij}$), activity (${Act}_{i}$) and occupation ($O_{i}$) of primary infection *i* are summarized in Table S4*.*

**Table S4. Hypothetical probability that the secondary infection** $\boldsymbol{j}$ **worked as a general worker, a service worker at the Xinfadi Market or a service worker in other places.**

| Condition | | Hypothetical probability^†^ | | |
| --- | --- | --- | --- | --- |
| $T_{ij}$ | ${Act}_{i}$ | $\psi_{j}^{GW}$ | $\psi_{j}^{XFD-SW}$ | $\psi_{j}^{Other-SW}$ |
| $O_{i}$ = GW |  |  |  |  |
| At home | / | 0.903 | 0 | 0.097 |
| In the workplace | / | 1 | 0 | 0 |
| In the community | Social activity | 0.903 | 0 | 0.097 |
| $O_{i}$ = XFD-SW |  |  |  |  |
| At home | / | 0.72 | 0.2 | 0.08 |
| In the community | Work | 0.4 | 0.2 | 0.4 |
| In the community | Social activity | 0.72 | 0.2 | 0.08 |
| $O_{i}$ = Other-SW |  |  |  |  |
| At home | / | 0.903 | 0 | 0.097 |
| In the community | Work | 0.8 | 0 | 0.2 |
| In the community | Social activity | 0.903 | 0 | 0.097 |
| $O_{i}$ = Nonworker |  |  |  |  |
| At home | / | 0.903 | 0 | 0.097 |
| In the community | Social activity | 0.903 | 0 | 0.097 |

Note: ^†^ The probabilities are hypothesized based on epidemiological investigations of the outbreak and the demographical structures in Beijing[7].

#### 3.4 Population interaction network based on spatially resolved mobility patterns

To emulate the spatial dispersion of SARS-CoV-2 infections, we first constructed the spatially resolved population mobility patterns in Beijing on the basis of the mobility data (see Section 1.2 for details) and then developed a network interaction model based on the constructed mobility patterns to stochastically assign the residential locations of each person with an infection, the workplace of each worker and the location for social activity if the transmission occurred through social contact. Details are described below.

##### 3.4.1 Spatially resolved mobility patterns in Beijing

The mobility data, provided by one of the leading mobile phone service providers in China, gave the total number of trips between each individual street/town by hour, stratified by the age and sex group of mobile phone users. By aggregating the mobility data, we obtained the average number of trips from the origin location $O$ to the destination location $D$ at time $t$ (in hours) in a given age and sex group on workday $n_{O\to D}^{wkd}(t|age, sex)$ and on holiday $n_{O\to D}^{hld}(t|age, sex)$, respectively. Assuming that the working-age population goes to work during morning rush hours (i.e., between 6:00 am and 10:00 am on workdays), we estimated the average daily trips from location $O$ to location $D$ for work ($n_{O\to D}^{work}$) as

$$n_{O\to D}^{work}=\sum_{t=6}^{10} n_{O\to D}^{wkd}(t|wk\_age)$$

where $wk\_age$ refers to working age groups (i.e., 18-59 years for males and 18-54 years for females). Since service workers provide social services to the general population, we assumed that their spatial mobility patterns for work were in accordance with and account for 10% of the population flows on holiday morning rush hours. Therefore, the average number of daily trips of the service workers from $O$ to $D$ for work $n_{O\to D}^{sw}$ is given by

$$n_{O\to D}^{sw}=0.1*\sum_{t=6}^{10} n_{O\to D}^{hld}(t|wk\_age)$$

and that of the general workers $n_{O\to D}^{gw}$ can be derived as

$n_{O\to D}^{gw}=n_{O\to D}^{work}-n_{O\to D}^{sw}$.

Trips at other times or of other age groups were considered social activities. The average daily trips from $O$ to $D$ for social activities on workday $n_{O\to D}^{soc\_wkd}$ and on holiday $n_{O\to D}^{soc\_hld}$ are given as

$$n_{O\to D}^{soc\_wkd}=\sum_{t} n_{O\to D}^{wkd}(t|all)-n_{O\to D}^{work}-n_{D\to O}^{work}$$

$$n_{O\to D}^{soc\_hld}=\sum_{t} n_{O\to D}^{hld}(t|all)-n_{O\to D}^{sw}-n_{D\to O}^{sw}$$

where $all$ refers to all age and sex groups, assuming that those who live in location $D$ and work at location $O$ would travel from $D$ to $O$ in the morning and back from $O$ to $D$ in the evening.

Then, the interaction matrices reflecting the population mobility patterns in Beijing in the absence of nonpharmaceutical interventions were constructed based on the aggregated data before the Xinfadi outbreak (June 1 to June 12, 2020), with each row of the matrices representing the origin street/town (i.e., residential location) and each column representing the destination street/town (i.e., location for work or social activity). Specifically, the interaction matrices for work are defined as $I^{gw}$ (for general workers) and $I^{sw}$ (for service workers), of which each cell represents the probability of working at location $D$ for an individual living in location $O$, given as $I_{O\to D}^{gw}=\frac{n_{O\to D}^{gw}}{\sum_{K} n_{O\to K}^{gw}}$ and $I_{O\to D}^{sw}=\frac{n_{O\to D}^{sw}}{\sum_{K} n_{O\to K}^{sw}}$. Similarly, the interaction matrices reflecting the social mobility patterns on workday ($I^{soc\_wkd}$) and on holiday ($I^{soc\_hld}$) are constructed as $I_{O\to D}^{soc\_wkd}=\frac{n_{O\to D}^{soc\_wkd}}{\sum_{K} n_{O\to K}^{soc\_wkd}}$ and $I_{O\to D}^{soc\_hld}=\frac{n_{O\to D}^{soc\_hld}}{\sum_{K} n_{O\to K}^{soc\_hld}}$, respectively.

##### 3.4.2 Network interaction model

Locations of secondary infections were stochastically allocated using a spatially structured network interaction model based on the constructed mobility patterns. We started with the people with initial infections, whose home addresses, workplaces (i.e., the Xinfadi Market in Huaxiang Street, Fengtai District, Beijing), and household sizes ($hh\_size$) were collected through epidemiological investigations. We layered their home address into 3 levels with incremental spatial accuracy: i) the street/town (the basic-level administrative divisions in China); ii) the block; and iii) the household. We created a unique block identification number $blk\_id$ for each residential block and a unique household identification number $hh\_id$ for each family, where residents of a block shared the same $blk\_id$ and members of a household shared the same $hh\_id$. Then, for any transmission between an initial infection and a secondary infection (and all subsequent infections), the residential location (i.e., the street/town, $blk\_id$ and $hh\_id$), the workplace (if he or she worked), the social place (if the transmission event occurred through social activity), and the household size ($hh\_size$) of the secondary infection were probabilistically allocated through a network interaction model based on their transmission setting.

- **Household transmission:** For any transmission from primary infection$i$ to secondary infection $j$ that occurred at home, we assumed that all interactions were with the members of the household; thus, $j$’s residential location and household size were the same as those of the primary infector $i$, i.e.,

${loc\_resid}_{j}={loc\_resid}_{i}$,

${blk\_id}_{j}={blk\_id}_{i}$,

${hh\_id}_{j}={hh\_id}_{i}$,

${hh\_size}_{j}={hh\_size}_{i}$.

The workplace of the secondary infection $j$ (${loc\_wk}_{j}$) is then chosen according to his or her occupation based on the mobility patterns for work in Beijing, given as

$P_{({loc\_wk}_{j}=D)}=I_{{loc\_resid}_{j}\to D}^{gw}$ (if $j$ is a general worker)

or

$P_{({loc\_wk}_{j}=D)}=I_{{loc\_resid}_{j}\to D}^{sw}$ (if $j$ is a service worker),

where $P_{({loc\_wk}_{j}=D)}$ indicates the probability that $j$ worked at location $D$, $I^{gw}$ and $I^{sw}$ are interaction matrices for work as defined in Section 3.4.1, while for any secondary infection who worked at the Xinfadi Market, the workplace is Huaxiang Street (where the Xinfadi Market is located).

- **Workplace transmission:** if a transmission event occurred in the workplace, we assumed that the primary infection$i$ and the secondary infection $j$ shared the same workplace, i.e.,

${loc\_wk}_{j}={loc\_wk}_{i}$.

The residential street/town of the secondary infection $j$ (${loc\_resid}_{j}$) was then allocated according to the mobility patterns, given as

$P_{({loc\_resid}_{j}=O)}=I_{O\to{loc\_wk}_{j}}^{gw}$,

where workplace transmission only occurs between general workers because service workers work in the community (as assumed in Section 3.3). We further randomly chose a $blk\_id$ and a $hh\_id$ for $j$ within his or her residential street/town. The household size of $j$ was stochastically generated based on the age-specific household size distribution (Table S5) collected through a survey study in China[6].

**Table S5. Age-specific household size distribution.**

| **Household size** | **Probability** | | |
| --- | --- | --- | --- |
|  | **0-18 years** | **19-64 years** | **≥65 years** |
| 1 | 0.005 | 0.037 | 0.070 |
| 2 | 0.000 | 0.239 | 0.573 |
| 3 | 0.525 | 0.517 | 0.171 |
| 4 | 0.190 | 0.105 | 0.090 |
| 5 | 0.253 | 0.086 | 0.070 |
| 6 | 0.023 | 0.017 | 0.010 |
| 7 | 0.005 | 0.000 | 0.010 |
| 8 | 0.000 | 0.000 | 0.005 |

- **Community transmission:** For any transmission that occurred in the community, we first allocated a transmission location (${loc}_{i\to j}^{trans}$) according to the activity of the primary infection $i$ ($\mathrm{Act}_{i}$, see Section 3.3 for definition). If the primary infection $i$ infected the secondary infection $j$ through work contact (i.e., $\mathrm{Act}_{i}=work$), we have

${loc}_{i\to j}^{trans}={loc\_wk}_{i}$,

Otherwise, if $i$ infected $j$ through social contact (i.e., $\mathrm{Act}_{i}=social$), we assumed 60% of his or her social activities were short distance travels within neighbourhoods, i.e., $P_{({travel\_in\_blk}_{i}=1)}^{soc}=0.6$. If ${travel\_in\_blk}_{i}=1$, we have

${loc}_{i\to j}^{trans}={loc\_resid}_{i}$,

If ${travel\_in\_blk}_{i}=0$, ${loc}_{i\to j}^{trans}$is stochastically generated following the probabilities given below:

$P_{({loc}_{i\to j}^{trans}=D)}=I_{{loc\_resid}_{i}\to D}^{soc\_wkd}$ (on a workday),

$P_{({loc}_{i\to j}^{trans}=D)}=I_{{loc\_resid}_{i}\to D}^{soc\_hld}$ (on a holiday),

where $P_{({loc}_{i\to j}^{trans}=D)}$ represents the probability that the transmission event occurred at location $D$. Then, the residential street/town (${loc\_resid}_{j}$) and the workplace (${loc\_wk}_{j}$) of the secondary infection $j$ are generated according to $\mathrm{Act}_{j}$ and ${loc}_{i\to j}^{trans}$: i) if the secondary infection $j$ became infected through work contact (i.e., $\mathrm{Act}_{j}=work$), we have

${loc\_wk}_{j}={loc}_{i\to j}^{trans}$.

We assume $P_{(travel\_in\_blk=1)}^{sw}=0.6$, where $P_{(travel\_in\_blk=1)}^{sw}$ indicates the probability of a service worker working near his or her residential block. If ${travel\_in\_blk}_{j}=1$, his or her residential location is given by

${loc\_resid}_{j}={loc\_wk}_{j}$,

Otherwise, it is randomly generated based on the population mobility patterns for service work in Beijing:

$P_{({loc\_resid}_{j}=O)}=I_{O\to{loc\_wk}_{j}}^{sw}$,

where $P_{({loc\_resid}_{j}=O)}$ represents the probability that $j$ lives in location $O$. ii) If the secondary infection $j$ became infected through social contact (i.e., $\mathrm{Act}_{j}=social$), we assume 60% of his or her social activities were short distance travels within neighbourhoods, i.e., $P_{({travel\_in\_blk}_{j}=1)}^{soc}=0.6$, which cannot be captured by mobile phone signalling data. If ${travel\_in\_blk}_{j}=1$, we have

${loc\_resid}_{j}={loc}_{i\to j}^{trans}$,

Otherwise, it is randomly generated based on the population mobility patterns for social activities in Beijing:

$P_{({loc\_resid}_{j}=O)}=I_{O\to{loc}_{i\to j}^{trans}}^{soc\_wkd}$ (on workday),

$P_{({loc\_resid}_{j}=O)}=I_{O\to{loc}_{i\to j}^{trans}}^{soc\_hld}$ (on holiday),

where $P_{({loc\_resid}_{j}=O)}$ represents the probability that $j$ lives in location $O$. The workplace of $j$ (${loc\_wk}_{j}$) is then chosen according to his or her occupation, given as

$P_{({loc\_wk}_{j}=D)}=I_{{loc\_resid}_{j}\to D}^{gw}$ (if $j$ is a general worker)

or

$P_{({loc\_wk}_{j}=D)}=I_{{loc\_resid}_{j}\to D}^{sw}$ (if $j$ is a service worker),

where $P_{({loc\_wk}_{j}=D)}$ indicates the probability that $j$ works at location $D$. If $j$ works at the Xinfadi Market, the workplace is Huaxiang Street. Next, we randomly chose a $blk\_id$ and a $hh\_id$ for $j$ within his or her residential street/town. If ${travel\_in\_blk}_{i}=1$ and ${travel\_in\_blk}_{j}=1$, indicating that the primary infection $i$ and the secondary infection $j$ live in the same block, we have ${blk\_id}_{j}={blk\_id}_{i}$. Finally, the household size of $j$ was stochastically generated based on the household size distribution in Table S5. However, if $j$ worked at the Xinfadi Market, his or her household size was derived from the household size distribution of Xinfadi workers collected from close contact data, as they are a special group of people mainly living in the staff dormitories near the market.

#### 3.5 Nonpharmaceutical interventions (NPIs)

3.5.1 **Symptomatic surveillance**

Outpatients at hospitals and local clinics with symptoms consistent with the clinical features of COVID-19 were immediately isolated as SARS-CoV-2 suspected cases until they were fully recovered and no longer infectious. Initially, we assumed that 33% of the people with symptomatic infections (${i.e., \Phi}_{hosp}=33\%$) would seek medical attention after a mean time delay of 3.7 days from the onset of symptoms, drawing from a Weibull distribution (shape = 2.38, scale = 4.17) derived from the epidemiological data. After the official report of the outbreak on June 13, 2020, with enhanced symptom surveillance in the community, we assumed that more people with symptomatic infections (${i.e., \Phi}_{hosp}=67\%$) would seek health care consultation after a shorter time delay with a mean of 2.7 days from symptom onset, drawing from a gamma distribution (shape = 0.69, rate = 0.26). Three RT‒PCR (i.e., reverse transcription polymerase chain reaction) tests for SARS-CoV-2 diagnosis were conducted on the 1^st^, 3^rd^ and 7^th^ days of isolation. Each testing time had to be completed (i.e., from collection of samples to reporting of results) within six hours. The sensitivity of RT‒PCR testing was assumed to vary with time, following the estimates of a prior study[8].

3.5.2 **Mask wearing**

Mask wearing was required in public spaces, but its implementation was affected by individual compliance. For each transmission event in the unmitigated transmission chains, we stochastically generated the mask-wearing status (i.e., whether an individual wore a mask at the time of transmission) of both the infector and the infectee, assuming 20% of the population wore masks in the workplace and 50% in the community. Then, the transmission could be probabilistically truncated by protection from masks. The protective effect of mask wearing against further transmission and infection of SARS-CoV-2 was assumed to be 9.5% and 18%, respectively[9, 10].

3.5.3 **Closure of the Xinfadi Market**

The Xinfadi Market was closed on June 13, 2020, which implies that workers in the market would no longer be infected or infect others through work. We thus removed the corresponding transmission events from the branching tree and truncated the subsequent transmission.

3.5.4 **Quarantine and testing of key populations**

The key population, defined as individuals associated with the Xinfadi Market, including workers at the Xinfadi Market, visitors to the Xinfadi Market and residents living around the Xinfadi Market, were tested and/or quarantined in the early phase of the outbreak. Detailed methods are briefly described below.

- **Workers at the Xinfadi Market:** Workers at the Xinfadi Market were assessed to be at the highest risk. They were quarantined in centralized facilities for medical observation for at least 14 days. We assumed that 40%, 40% and 20% of the workers were quarantined on June 12^th^, 13^th^, and 14^th^, 2020, respectively. Periodic RT‒PCR testing was conducted on the 1^st^, 4^th^, 7^th^ and 14^th^ days of quarantine and the 2^nd^ and 7^th^ days after discharge. For workers who developed symptoms during the quarantine period, additional tests were conducted on the 1^st^, 3^rd^, and 7^th^ days after symptom onset. Individuals diagnosed by RT‒PCR testing were immediately transported to designated hospitals for treatment until they fully recovered.
- **Visitors to the Xinfadi Market:** Visitors who had been to the Xinfadi Market between May 30 and June 12, 2020, were asked to stay at home for 14 days. We assumed that 20%, 30%, 30% and 20% of the visitors were traced and quarantined on June 12^th^, 13^th^, 14^th^ and 15^th^, 2020, respectively. Home quarantine is assumed to be completely effective in preventing workplace and community transmission but cannot prevent household transmission. RT‒PCR testing was performed on the 1^st^, 7^th^ and 14^th^ days of home quarantine, and people who tested positive were immediately taken to designated hospitals for treatment.
- **Residents living around the Xinfadi Market:** Residents living around the Xinfadi Market were confined to their living communities from June 13, 2020, until no new infections were reported for 14 consecutive days. Mass testing was implemented every seven days during the confinement period. People who tested positive were taken to designated hospitals for treatment.
  3.5.5 **Contact tracing**

Contact tracing starts at the time of laboratory confirmation of an index case. Close contact was defined as a person who interacted with a confirmed or suspected COVID-19 case from 4 days before to 14 days after illness onset or with an asymptomatic carrier from 4 days before to 14 days after collection of the first positive sample. Close contacts were further grouped into household contacts, work contacts and community contacts based on their transmission setting. We assumed that all household contacts were immediately quarantined (i.e., 50% on the same day of index case diagnosis and 50% on the next day), while all work contacts and 70% of the community contacts quarantined with a time delay (mean = 3.0 days) following a gamma distribution (shape = 1.79, rate = 0.60) derived from the contact tracing data. Centralized quarantine at designated facilities for at least 14 days was required for all close contacts, with periodic RT‒PCR testing on the 1^st^, 4^th^, 7^th^ and 14^th^ days of quarantine and the 2^nd^ and 7^th^ days after discharge. Assuming centralized quarantine was completely effective in preventing further transmission, we thus removed all further transmission after an infected individual was quarantined as a close contact.

3.5.6 **Residential community confinement**

Since June 13, 2020, residential communities with detected infections have been on lockdown at the block level until 14 days after the identification of the last case, with stay-at-home orders for all residents other than essential workers. We thus removed all transmission that required moving in or out of the confined communities during the lockdown period. Since community workers need to provide basic living supplies for confined residents, we stochastically retained 10% of the transmission within the residential blocks.

3.5.7 **Mobility restrictions**

Prior to June 11, 2020, Beijing had reported no new infections for 56 consecutive days, therefore allowing unrestricted movement. After the Xinfadi outbreak, the street/town was upgraded to moderate risk once it had reported more than one infection and then upgraded to high risk when more than 5 infections were reported, while other regions, without or with one detected infection, remained low-risk areas. Population mobility restrictions were implemented in risk regions, with entertainment venues being closed, mass gatherings being prohibited, and/or unnecessary travel being banned. We quantified the reduction in population flows based on mobile phone signalling data, as shown in Fig. S3. For high-risk regions (Fig. S3A), taking Huaxiang Street as an example, we observed that the trips entering, leaving and moving within the streets dropped to 30% of pre-epidemic levels. For moderate-risk areas (Fig. S3B), such as Xiluoyuan Street, the population flows moving in and out of the street were reduced by approximately 50%, while short-distance travel within the streets was reduced by approximately 40%. For low-risk regions (Fig. S3C), such as Jiuxianqiao Street, unrestricted movement was allowed. However, we observed approximately 20% and 10% reductions in the population flows moving in/out and within the streets of low-risk areas, respectively, which may be due to changes in human behaviours after the outbreak. The hypothetical origin-destination mobility matrix depending on risk levels is constructed and shown in Table S6. We then stochastically removed the transmission events from the unmitigated transmission chains accordingly.

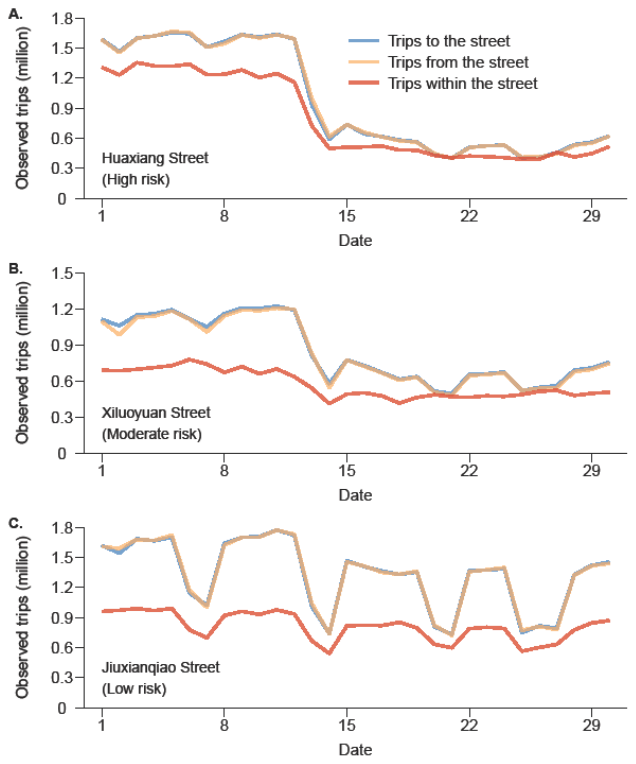

**Figure S3. Observed trips moving in and out and within areas by risk levels** based on mobile phone signalling data. **A**. High-risk region (Huaxiang Street). **B**. Moderate-risk region (Xiluoyuan Street). **C**. Low-risk region (Jiuxianqiao Street).

**Table S6. The hypothetical origin-destination mobility matrix depending on risk levels.**

| Risk level of the origin street/town | Risk level of the destination street/town | | | Mobility within one street/town |
| --- | --- | --- | --- | --- |
|  | High | Moderate | Low |  |
| High | 0 | 0.1 | 0.3 | 0.3 |
| Moderate | 0.1 | 0.3 | 0.5 | 0.6 |
| Low | 0.3 | 0.5 | 0.8 | 0.9 |

Note: The risk level refers to the real-time risk level of the street/town at the time of transmission. The value in each cell of the matrix refers to the average travel probability per person after the Xinfadi outbreak, given the risk level of the origin and destination regions.

3.5.8 **Mass testing**

On June 13, 2020, a population-wide mass testing programme was initiated in Beijing. Three rounds of RT‒PCR testing was required for all residents living in streets/towns with detected infections, with each round of mass testing being completed within 3-4 days. The interval between each round of testing was usually 7 days. Residents who tested positive were immediately isolated for treatment or medical observation.

### 4 Increasing transmissibility and immune evasion properties of SARS-CoV-2 variants

In Fig. 1A, we schematically plotted the relative transmissibility and immune evasion properties of SARS-CoV-2 variants compared to the ancestral strain based on estimates in prior studies[11-16]. The solid lines and the arrows indicate the evolutionary trajectory of SARS-CoV-2, suggesting strong adaptive evolution of SARS-CoV-2 in the direction of increasing transmissibility and immune evasion properties.

### 5 Controllability of the SARS-CoV-2 ancestral strain across the globe

As of mid-2020, the COVID-19 pandemic, caused by the ancestral SARS-CoV-2 strain, has spread across the world. Using the 14-day smoothing epidemic/death curves publicly available at Johns Hopkins University Center for Systems Science and Engineering (JHU CCSE) coronavirus website, we estimated the time-varying effective reproduction numbers (*R_t_*) during the first COVID-19 wave in different countries/states (as listed in Table S7) with high-quality surveillance data based on the method proposed by Cori *et al.*[17]. We found that due to the limited transmissibility of the ancestral strain (*R_0_*=2.5[2]), multiple countries/states with different socioeconomic statuses successfully achieved temporal eradication of SARS-CoV-2 in the early stage of the pandemic (i.e., with *R_t_* decreasing below the epidemic threshold of 1) through implementing NPIs even though population immunity was neglectable (i.e., no effective vaccine was available) (Fig. 1B).

**Table S7. The data sources and countries/states of each region shown in Fig. 1B.**

| Region | Country | State | Source data |
| --- | --- | --- | --- |
| African Region | Algeria | - | Epidemic curve |
| Eastern Mediterranean Region | Afghanistan | - | Epidemic curve |
| Eastern Mediterranean Region | Pakistan | - | Death curve |
| Eastern Mediterranean Region | Sudan | - | Death curve |
| European Region | France | - | Death curve |
| European Region | Germany | - | Epidemic curve |
| European Region | Ireland | - | Epidemic curve |
| European Region | Spain | - | Epidemic curve |
| European Region | United Kingdom | - | Epidemic curve |
| Southeast Asian Region | Thailand | - | Epidemic curve |
| Southeast Asian Region | Bangladesh | - | Death curve |
| Western Pacific Region | Japan | - | Epidemic curve |
| Western Pacific Region | Korea, South | - | Epidemic curve |
| Western Pacific Region | New Zealand | - | Epidemic curve |
| Western Pacific Region | Singapore | - | Epidemic curve |
| Region of the Americas | Canada | - | Death curve |
| Region of the Americas | Chile | - | Epidemic curve |
| Region of the Americas | US | Connecticut | Epidemic curve |
| Region of the Americas | US | Columbia | Death curve |
| Region of the Americas | US | Illinois | Death curve |
| Region of the Americas | US | Indiana | Death curve |
| Region of the Americas | US | Massachusetts | Death curve |
